# Prevalence and Predictors of the Double Burden of Malnutrition among Under-Five Children in Urban Ghana

**DOI:** 10.1101/2025.08.01.25332623

**Authors:** Deborah Balapou, John Foster Atta-Doku, Wisdom Kwaku Achiam Amuka, Richard Osei Agjei, Emmanuel Kumah

**Affiliations:** University of Education, Winneba, Faculty of Health, Allied Sciences and Home Economics Education, Department of Health Administration and Education P. O. Box 25., Winneba, Central Region, Ghana, West Africa

**Author notes:** Corresponding Author: John Foster Atta-Doku.

**Keywords:** Double burden of malnutrition, undernutrition, overnutrition, under-five children, child nutrition, breastfeeding practices

## Abstract

**Introduction:** The double burden of malnutrition, defined as the coexistence of undernutrition and overnutrition within the same individual, is an emerging public health challenge in low- and middle-income countries undergoing rapid urbanization. In Ghana, the rising prevalence among under-five children underscores the need for context-specific evidence. This study examined the prevalence and predictors of the double burden of malnutrition among under-five children in a selected urban area in Ghana.

**Methods:** A hospital-based cross-sectional study was conducted among 271 mother-child pairs using a semi-structured questionnaire and standardized anthropometric measurements. Nutritional status was assessed using the World Health Organization’s Child Growth Standards. The double burden of malnutrition was defined as the co-occurrence of underweight or overweight with any form of stunting within the same child. Descriptive statistics, Chi-square tests, and binary logistic regression were used to analyze the data.

**Results:** The prevalence of the double burden of malnutrition among the sampled children was 31.37%. Significant predictors included child’s age (OR=1.034; p=0.002), low physical activity levels (OR=6.22; p=0.001 for inactive children), lack of breastfeeding (OR=11.82; p=0.001), non-exclusive breastfeeding (OR=6.06; p=0.001), absence of formula feeding (OR=2.16; p=0.043), and early introduction of semi-solid foods (OR=0.28; p=0.019 for introduction at 6–8 months versus <6 months). Additionally, maternal tertiary education was protective against the double burden of malnutrition (OR=0.24; p=0.032).

**Conclusion:** The findings highlight the multifactorial nature of DBM in urban Ghana, shaped by child feeding practices, physical activity, and maternal education. Addressing DBM will require integrated public health strategies that promote optimal infant feeding, physical activity, and maternal health education, particularly in urban poor communities.

## Introduction

Malnutrition remains a significant global health concern, particularly in low- and middle-income countries (LMICs), where both undernutrition and overnutrition are increasingly co-occurring [1]. Traditionally burdened by child stunting, wasting, and micronutrient deficiencies [2,3], many LMICs now face rising rates of overweight, obesity, and diet-related non-communicable diseases [2]. The concurrent presence of these conditions within the same population, individual, or setting is described as the double burden of malnutrition (DBM) [4,5]. This dual manifestation reflects persistent inequities in food systems, access to health, and socioeconomic development, presenting complex challenges for public health systems undergoing rapid nutritional and demographic transitions [1].

Although there is no single standardized definition of DBM, it typically includes combinations of undernutrition (such as stunting or wasting) and overnutrition (such as overweight or obesity). This complexity contributes to inconsistencies across studies [6], yet there is broad consensus on the need for integrated interventions that can simultaneously address both forms of malnutrition. Urbanization plays a key role in shaping dietary practices and physical activity patterns in LMICs [7,8]. Urban environments often expose populations to processed, energy-dense foods while limiting access to affordable, nutrient-rich options [9]. At the same time, physical inactivity and uneven access to healthcare, sanitation, and education further compound nutritional vulnerabilities [9,10].

In sub-Saharan Africa (SSA), this shifting landscape has led to a rise in child-level forms of DBM, where children may experience coexisting indicators such as underweight, stunting, and overweight [5,11]. These patterns are driven by structural factors including poverty, food insecurity, suboptimal infant feeding practices, and disparities in maternal education and employment [12,13]. Although maternal overweight and child undernutrition within the same household have received attention in the literature, child-centered manifestations of the double burden of malnutrition are more prevalent and require context-specific evidence to guide effective interventions [14].

Rapid urbanization and dietary shifts have led to simultaneous gains in child survival and rising overweight among both children and adults in Ghana [15,16]. However, these trends have been unevenly distributed. National and subnational surveys indicate that undernutrition persists among urban poor populations, despite the emergence of overweight and obesity among children with increased access to calorie-dense foods and reduced physical activity [17,18]. These coexisting trends highlight the need for targeted, equity-focused research into the social and behavioral drivers of child malnutrition.

Despite growing awareness of DBM, there remains limited empirical evidence on its prevalence and determinants among children under five in Ghana’s urban settings. This study investigates the double burden of malnutrition among under-five children in Ghana, focusing specifically on its prevalence and associations with child-level feeding practices, physical activity, and maternal socio-demographic characteristics. It is guided by the United Nations Children’s Fund (UNICEF) conceptual framework on the determinants of child malnutrition, which recognizes the influence of immediate, underlying, and basic causes of malnutrition across individual, household, and structural levels [19,20,21]. The framework informs the selection and interpretation of variables in the analysis and supports a more holistic understanding of the multifaceted drivers of DBM. The findings aim to inform public health strategies that acknowledge the complexity of child malnutrition and support the design of integrated interventions in urban Ghana.

## Methods

### Study Area and Institution

The study was conducted in Kumasi, the capital of Ghana’s Ashanti Region and the country’s second-largest urban center. Situated about 200 kilometers northwest of Accra, Kumasi spans roughly 214 square kilometers in the tropical forest belt. The 2021 census recorded around 443,981 residents in the core city, while the Greater Kumasi area exceeded 3 million [22]. Once known as the “Garden City of West Africa,” Kumasi has seen rapid urbanization and related planning challenges [23].

Komfo Anokye Teaching Hospital (KATH), located in Kumasi, served as the study site. Established in 1954 and affiliated with the Kwame Nkrumah University of Science and Technology (KNUST), KATH is Ghana’s second-largest hospital and a key referral center for the Ashanti, Bono, Ahafo, and northern regions. The hospital provides specialized maternal and child health services and hosts a large, diverse patient population [24,25], making it an appropriate setting to investigate child malnutrition.

### Study Design and Sampling Procedure

A hospital-based cross-sectional study design was employed, using quantitative methods. The target population consisted of children aged 0–59 months and their mothers aged 15–49 years attending outpatient services at KATH. Participants were selected using convenience sampling. Mothers outside the specified age range, children over five years, and non-maternal caregivers were excluded.

Based on hospital records, an estimated 56 children visited the facility daily. Over the 15-day data collection period, the study population was projected at 840 (56 × 15) mother-child pairs.

The sample size was determined using the Taro Yamane formula: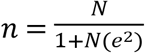

Where:

n = required sample size

N = population size (840)

e = margin of error (0.05 for 95% confidence level)

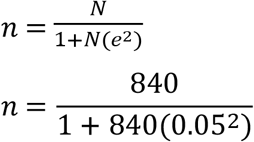

n = 270.9 ∼ 271

### Data Collection Instruments

Data were collected using two main instruments developed by the researchers: a semi-structured questionnaire and standardized anthropometric tools. The questionnaire was adapted from validated instruments used in earlier studies [26,27] and modified to suit the study context. It included sections on socio-demographic characteristics, obstetric history, environmental factors, infant feeding practices, and dietary behaviors.

Anthropometric data were collected using calibrated equipment. A digital Seca scale was used for weighing children under two years, while a standard adult scale was used for older children and mothers. Lengths of children under two were measured using a recumbent infantometer, and heights of older children and mothers were measured with a stadiometer or measuring tape. All measurements were recorded to the nearest 0.1 kg or 0.1 cm. Daily calibration and zeroing procedures were conducted to ensure reliability.

### Assessment of Nutritional Status

Children’s nutritional status was assessed using the 2006 WHO Child Growth Standards. Weight-for-age, height-for-age, and weight-for-height Z-scores were generated using the WHO Anthro package in R. Children with Z-scores below -2 standard deviations from the median were classified as undernourished. Based on WHO classifications, BMI values were categorized into underweight, normal weight, overweight, and obese.

In this study, DBM was defined as the coexistence of stunting (moderate or severe) alongside either overweight/obesity or underweight within the same child. Although conventional definitions of DBM emphasize the co-occurrence of undernutrition and overnutrition, especially overweight with stunting, this study captured both underweight–stunting and overweight–stunting combinations to represent the full spectrum of nutritional challenges observed in urban Ghana. This broader operationalization reflects emerging evidence on compounded malnutrition forms that impair child development in resource-limited settings [28,29].

### Validity of Data Collection Instruments

Content validity was addressed through a review of the questionnaire and research protocol by a public health expert and the Research Department at KATH. Revisions were made based on expert feedback to enhance contextual appropriateness and clarity. A pilot test was conducted among a small sample of mother-child pairs to assess question sequence, comprehension, and flow.

### Data Collection

Data collection commenced after obtaining ethical approval from the KATH Institutional Review Board and administrative clearance from hospital authorities. Written informed consent was obtained from all participating mothers, including consent from a parent or guardian for participants under 18 years of age. Data were collected over 15 consecutive working days from 19th June to 7th July 2023 at the hospital’s outpatient department. Trained research assistants administered the questionnaires and performed anthropometric measurements under daily supervision to ensure adherence to study protocols. Completed questionnaires were reviewed each day for accuracy and completeness before data entry and analysis.

### Data Analysis

Data analysis was conducted using R version 4.5.0 and STATA version 18. Descriptive statistics were used to summarize socio-demographic and nutritional characteristics. The prevalence of the double burden of malnutrition (DBM) was computed as the proportion of children exhibiting coexisting indicators of undernutrition (e.g., stunting or underweight) and overnutrition (e.g., overweight or obesity). Chi-square tests were used to explore bivariate associations between DBM and child/maternal characteristics. Variables with a p-value less than 0.25 in the bivariate analysis were included in a binary logistic regression model to identify independent predictors of DBM. All predictor variables were checked for multicollinearity using Variance Inflation Factors (VIF) before the regression analysis. The results indicated no multicollinearity concerns, as all VIF values were within acceptable thresholds (see supplementary file). Odds ratios and confidence intervals were calculated and presented in tables to support interpretation.

## Results

### Children sociodemographic characteristics

A total of 271 under-five children were recruited at Komfo Anokye Teaching Hospital. The mean age was 20.63 months (SD ±1.08), with the largest group being those under six months (26.94%). Males comprised 54.98% of the sample. Most children were from Christian households (84.87%), and over half (56.09%) were reported as very active. Regarding fathers’ education, the majority had either secondary (36.53%) or tertiary education (35.79%). Please refer to Table 1 for more details.

**Table 1:** Children’s socio-demographic characteristics.

| *Child Socio-demographic Characteristics* | *Frequency (n)* | *Percentage (%)* |
| --- | --- | --- |
| ***Child's Age Category*** | Mean age in Months: 20.63 ± 1.08 |  |
| <6 Months | 73 | 26.94 |
| 6–12 Months | 55 | 20.30 |
| 13–24 Months | 61 | 22.51 |
| 25–36 Months | 36 | 13.28 |
| 37–59 Months | 46 | 16.97 |
| ***Child's Sex*** |  |  |
| Male | 149 | 54.98 |
| Female | 122 | 45.02 |
| ***Child's Religion*** |  |  |
| Christian | 230 | 84.87 |
| Islam | 40 | 14.76 |
| Traditional | 1 | 0.37 |
| ***Child's Birth Order*** |  |  |
| 1st | 75 | 27.68 |
| 2nd–4th | 166 | 61.25 |
| 5th–7th | 30 | 11.07 |
| ***Child's Daily Activities*** |  |  |
| Very Active | 152 | 56.09 |
| Active some of the time | 88 | 32.47 |
| Not Active | 31 | 11.44 |
| ***Father's Educational Level*** |  |  |
| No Formal Education | 12 | 4.43 |
| Basic | 56 | 20.66 |
| Secondary | 99 | 36.53 |
| Postsecondary | 7 | 2.58 |
| Tertiary | 97 | 35.79 |

### Children dietary practices

Table 2 presents the feeding practices of the children. The vast majority (96.31%) had ever been breastfed. Breastfeeding was initiated within six hours of birth in 55.72% of cases, with 39.48% initiating within the first hour and 4.80% after six hours. Only 40.59% of the children were exclusively breastfed for six months. Formula feeding was common; 70.11% received formula, with 49.47% of those introduced before six months. Regarding semi-solid food introduction, 53.14% began between six and eight months, 41.33% before six months, and 5.54% after eight months.

**Table 2:** Children’s dietary practices.

| **Variable** | **Frequency (n)** | **Percentage (%)** |
| --- | --- | --- |
| ***Breastfeeding (Ever)*** |  |  |
| Yes | 261 | 96.31 |
| No | 10 | 3.69 |
| ***Initiation of Breastfeeding*** |  |  |
| < One Hour | 107 | 39.48 |
| > 6 Hours | 13 | 4.80 |
| < 6 Hours | 151 | 55.72 |
| ***Exclusive Breastfeeding (6 Months)*** |  |  |
| Yes | 110 | 40.59 |
| No | 161 | 59.41 |
| ***Using Formula Feed*** |  |  |
| Yes | 190 | 70.11 |
| No | 81 | 29.89 |
| ***Introduced formula feed before six months (n=190)*** |  |  |
| Yes | 94 | 49.47 |
| No | 96 | 50.53 |
| ***Age of Semi-solid Food Introduction*** |  |  |
| < 6 Months | 112 | 41.33 |
| 6-8 Months | 144 | 53.14 |
| > 8 Months | 15 | 5.54 |

### Mothers’ sociodemographic characteristics

The sociodemographic profile of the mothers is presented in Table 3. The majority were aged 31–39 years (53.14%), followed by 20–30 years (30.63%), 40 years and above (14.39%), and below 20 years (1.85%). Most mothers were married (84.13%) and resided in urban areas (89.30%). Regarding education, 35.79% had tertiary education, 32.84% had basic education, 25.09% had secondary education, and a smaller number had postsecondary (1.11%) or no formal education (5.17%). In terms of employment, 80.07% were employed. Physical activity levels were low, with 84.50% reporting no weekly exercise, 14.39% exercising one to three days per week, and only 1.11% exercising more than four days weekly.

**Table 3:**
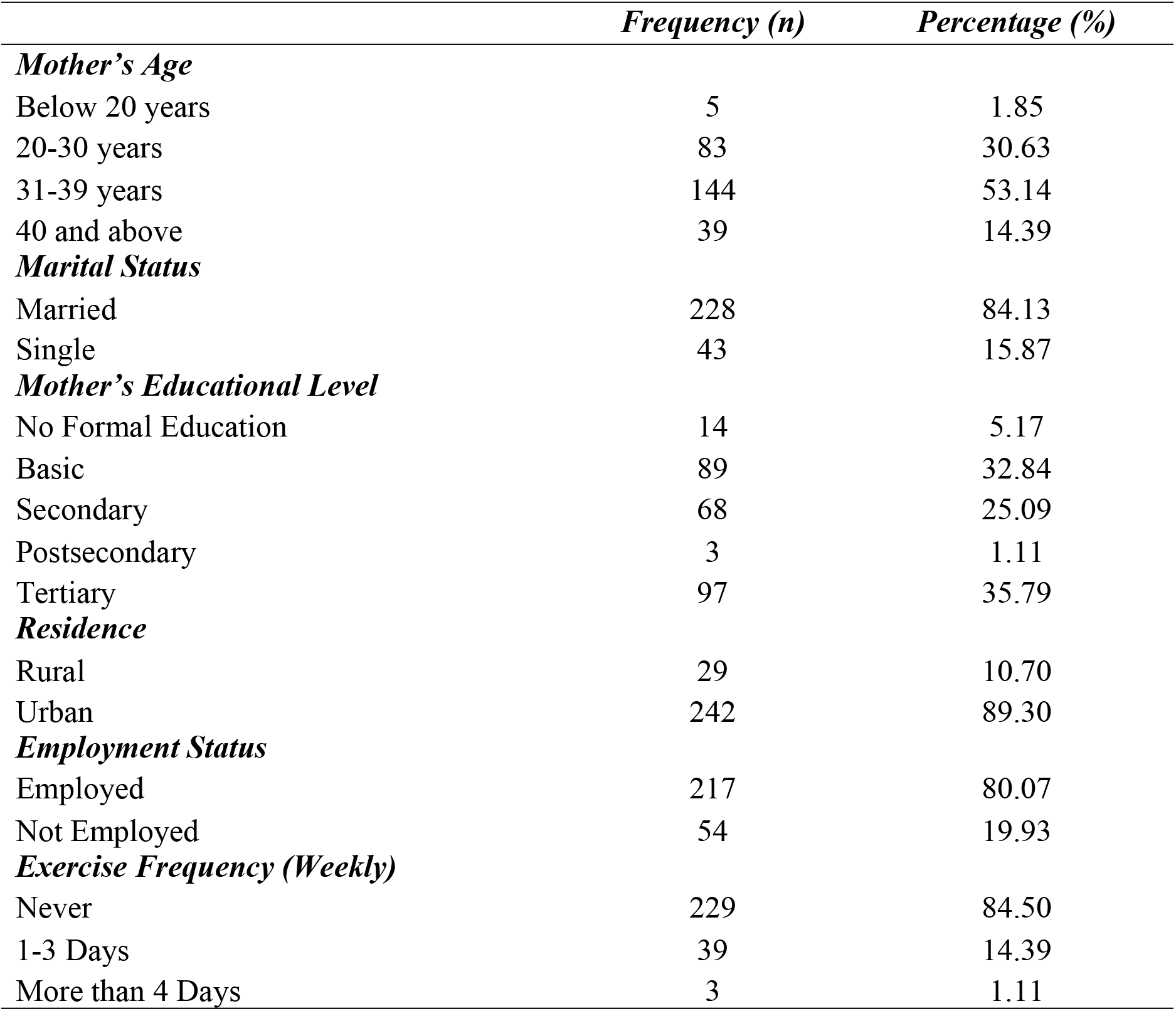
Mothers’ Socio-demographic characteristics.

### Nutritional Status of Children

Table 4 presents the nutritional classifications. The majority of children (82.29%) were underweight, 13.28% had normal BMI, and 4.43% were overweight or obese. For height-for-age, 63.10% were classified as normal, 19.93% as stunted, and 16.97% as severely stunted. Based on the study’s definition, the double burden of malnutrition (underweight–stunting or overweight–stunting) was identified in 31.37% of the children. Given the relatively low proportion of overweight children, it is likely that a significant portion of these cases involved underweight coexisting with stunting.

**Table 4:** Distribution of Children by BMI, Stunting, and Double Burden of Malnutrition.

|  | Frequency (n) | Percentage (%) |
| --- | --- | --- |
| ***BMI Category*** |  |  |
| Normal | 36 | 13.28 |
| Underweight | 223 | 82.29 |
| Overweight/Obese | 12 | 4.43 |
| ***Stunting Category*** |  |  |
| Normal | 171 | 63.10 |
| Stunted | 54 | 19.93 |
| Severely Stunted | 46 | 16.97 |
| ***Double Burden of Malnutrition*** |  |  |
| Yes | 85 | 31.37 |
| No | 186 | 68.63 |

### Bivariate Associations with the Double Burden of Malnutrition

Chi-square tests were conducted to examine associations between the double burden of malnutrition (DBM) and various child and maternal characteristics (Table 5). A significant association was found between DBM and child’s activity level (χ^2^ = 9.374, p = 0.009), breastfeeding history (χ^2^ = 3.955, p = 0.047), and use of formula feed (χ^2^ = 7.529, p = 0.006). Mother’s education was also significantly associated with DBM (χ^2^ = 11.846, p = 0.019). Other factors, including the child’s sex, birth order, maternal age, marital status, residence, employment, and physical activity, were not significantly associated.

**Table 5:** Association Between DBM and Child/Maternal Characteristics.

| | Double Burden<br>malnutrition | | | $\chi^2$ | p-value |
| --- | --- | --- | --- | --- | --- |
|  | Yes | No | Total |  |  |
| **Child's Sex** |  |  |  |  |  |
| Male | 101 | 48 | 149 | 0.111 | 0.739 |
| Female | 85 | 37 | 122 |  |  |
| **Birth Order** |  |  |  |  |  |
| 1 | 49 | 26 | 75 | 0.719 | 0.698 |
| 2-4 | 115 | 51 | 166 |  |  |
| 5-7 | 22 | 8 | 30 |  |  |
| **Child's Daily Activities** |  |  |  |  |  |
| Very Active | 114 | 38 | 152 | 9.374 | 0.009*** |
| Active sometimes | 57 | 31 | 88 |  |  |
| Not Active | 15 | 16 | 31 |  |  |
| **Father's Education** |  |  |  |  |  |
| No formal | 8 | 4 | 12 | 4.986 | 0.289 |
| Basic | 34 | 22 | 56 |  |  |
| Secondary | 65 | 34 | 99 |  |  |
| Postsecondary | 6 | 1 | 7 |  |  |
| Tertiary | 73 | 24 | 97 |  |  |
| **Breastfed Ever** |  |  |  |  |  |
| Yes | 182 | 79 | 261 | 3.955 | 0.047** |
| No | 4 | 6 | 10 |  |  |
| **Initiation of Breastfeeding** |  |  |  |  |  |
| < 1 hour | 71 | 36 | 107 | 2.164 | 0.339 |
| < 6 hours | 108 | 43 | 151 |  |  |
| > 6 hours | 7 | 6 | 13 |  |  |
| **Exclusive Breastfeeding (6 Months)** |  |  |  |  |  |
| Yes | 82 | 28 | 110 | 3.005 | 0.083* |
| No | 104 | 57 | 161 |  |  |
| **Have you/will you introduce formula feed?** |  |  |  |  |  |
| Yes | 140 | 50 | 190 | 7.529 | 0.006*** |
| No | 46 | 35 | 81 |  |  |
| **Introduced formula feed before 6 months (n = 190)** |  |  |  |  |  |
| Yes | 66 | 28 | 94 | 1.156 | 0.282 |
| No | 74 | 22 | 96 |  |  |
| **Age of Semi-solid Food Introduction** |  |  |  |  |  |
| < 6 months | 74 | 38 | 112 | 2.834 | 0.242 |
| 6-8 months | 104 | 40 | 144 |  |  |
| > 8 months | 8 | 7 | 15 |  |  |
| **Mother's Age** |  |  |  |  |  |
| < 20 yrs | 2 | 3 | 5 | 2.126 | 0.547 |
| 20-30 yrs | 57 | 26 | 83 |  |  |
| 31-39 yrs | 101 | 43 | 144 |  |  |
| 40+ yrs | 26 | 13 | 39 |  |  |
| **Marital Status** |  |  |  |  |  |
| Married | 161 | 67 | 228 | 2.615 | 0.106 |
| Single | 25 | 18 | 43 |  |  |
| **Mother's Education** |  |  |  |  |  |
| No formal | 6 | 8 | 14 | 11.846 | 0.019*** |
| Basic | 63 | 26 | 89 |  |  |
| Secondary | 41 | 27 | 68 |  |  |
| Postsecondary | 1 | 2 | 3 |  |  |
| Tertiary | 75 | 22 | 97 |  |  |
| **Residence** |  |  |  |  |  |
| Rural | 19 | 10 | 29 | 0.147 | 0.702 |
| Urban | 167 | 75 | 242 |  |  |
| **Employment** |  |  |  |  |  |
| Yes | 153 | 64 | 217 | 1.773 | 0.183 |
| No | 33 | 21 | 54 |  |  |
| **Exercise (Weekly)** |  |  |  |  |  |
| Never | 158 | 71 | 229 | 1.757 | 0.415 |
| 1-3 days | 27 | 12 | 39 |  |  |
| > 4 days | 1 | 2 | 3 |  |  |

\*\*\* $p < .01$ , \*\* $p < .05$ , \* $p < .1$

### Multivariate Predictors of Double Burden of Malnutrition

Binary logistic regression analysis was conducted to identify independent predictors of DBM (Table 6). Child’s age was positively associated with DBM (OR = 1.034; p = 0.002). Compared to very active children, those active only sometimes had 2.26 times higher odds of DBM (p = 0.039), while inactive children had 6.22 times higher odds (p = 0.001). Children who had never been breastfed were 11.82 times more likely to experience DBM (p = 0.001), and those not exclusively breastfed had 6.06 times higher odds (p = 0.001). Formula non-users had 2.16 times greater odds of DBM (p = 0.043). The introduction of semi-solid feed between six and eight months was associated with significantly lower odds compared to introduction before six months (OR = 0.279; p = 0.019. Regarding maternal factors, tertiary education was associated with significantly reduced odds of DBM compared to no formal education (OR = 0.243; p = 0.032). Basic education showed a marginal association (OR = 0.030; p = 0.065). Other maternal variables, including employment and marital status, were not significantly associated.

**Table 6:** Predictors of Double Burden of Malnutrition Among Under-Five Children.

| *Predictors* | *Odds ratio* | *St.Err.* | *p-value* | *95% CI* |  |
| --- | --- | --- | --- | --- | --- |
|  |  |  |  | *Low* | *High* |
| *Age in months* | 1.034 | 0.011 | 0.002*** | 1.013 | 1.056 |
| *Child's daily activity* |  |  |  |  |  |
| Very active <sup>b</sup> | - |  |  |  |  |
| Active some of the time | 2.262 | 0.895 | 0.039** | 1.041 | 4.914 |
| Not Active | 6.221 | 3.543 | 0.001*** | 2.038 | 18.995 |
| *Have/are you breastfeeding* |  |  |  |  |  |
| Yes <sup>b</sup> | - |  |  |  |  |
| No | 11.815 | 8.925 | 0.001*** | 2.688 | 51.934 |
| *Exclusive breastfeeding for six months.* |  |  |  |  |  |
| Yes <sup>b</sup> | - |  |  |  |  |
| No | 6.063 | 3.122 | 0.001*** | 2.21 | 16.634 |
| *Using formula feed* |  |  |  |  |  |
| Yes <sup>b</sup> | - |  |  |  |  |
| No | 2.162 | 0.822 | 0.043** | 1.026 | 4.555 |
| *Age of introducing semi-solid feed* |  |  |  |  |  |
| < 6 months <sup>b</sup> | - |  |  |  |  |
| 6-8 months | 0.279 | 0.152 | 0.019** | .096 | 0.81 |
| greater than 8 | 0.595 | 0.45 | 0.493 | 0.135 | 2.619 |
| *Marital status* |  |  |  |  |  |
| Married <sup>b</sup> | - |  |  |  |  |
| Single | 1.479 | 0.598 | 0.333 | 0.669 | 3.269 |
| *Educational level* |  |  |  |  |  |
| No formal education <sup>b</sup> | - |  |  |  |  |
| Basic | 0.030 | 0.196 | 0.065* | 0.084 | 1.078 |
| Secondary | 0.445 | 0.293 | 0.218 | 0.122 | 1.615 |
| Postsecondary | 1.004 | 1.496 | 0.998 | .054 | 18.646 |
| Tertiary | 0.243 | 0.160 | 0.032** | 0.067 | 0.886 |
| *Are you employed?* |  |  |  |  |  |
| Yes <sup>b</sup> | - |  |  |  |  |
| No | 1.593 | 0.583 | 0.204 | 0.777 | 3.265 |

\*\*\* $p < .01$ , \*\* $p < .05$ , \* $p < .1$ , <sup>b</sup> = Reference category

## Discussion

The coexistence of undernutrition and overnutrition observed in this study reflects broader nutritional transitions seen in urbanizing low- and middle-income countries, where rising overweight trends increasingly accompany persistent stunting and wasting [1,30]. Despite a relatively low proportion of overweight or obese children, the prevalence of DBM was considerably higher, suggesting that many cases involved underweight–stunting combinations. This pattern points to a unique form of nutritional duality, shaped more by chronic deprivation than dietary excess. Clarifying these underlying patterns through disaggregated DBM classifications is essential for identifying context-specific vulnerabilities and developing more precise, responsive interventions.

Physical activity emerged as a critical behavioral factor associated with the double burden of malnutrition. Children who were reported to be inactive or only intermittently active were significantly more likely to exhibit coexisting indicators of under- and overnutrition. The pattern reflects broader urban realities in Ghana and other sub-Saharan African settings, where safe, accessible play spaces are limited, and children’s routines are increasingly shaped by sedentary behaviors such as screen time. In environments characterized by spatial constraints and safety concerns, opportunities for regular movement and physical engagement may be compromised, contributing to poor nutritional outcomes [16,31]. These findings highlight the importance of child-centered urban planning and interventions that encourage daily physical activity.

Feeding practices were also strongly associated with DBM. Children who had never been breastfed or were not exclusively breastfed for the first six months showed higher odds of DBM. Exclusive breastfeeding supports healthy metabolic development, enhances immune function, and is protective against both stunting and excess weight gain [32,33]. The findings suggest that disruptions in breastfeeding, whether due to lack of maternal knowledge, work constraints, or sociocultural practices, may increase vulnerability to malnutrition in both forms.

An unexpected finding was that children who had not received formula feeding were significantly more likely to experience DBM. Exclusive breastfeeding remains the gold standard for infant nutrition; however, this result may reflect situations where breastfeeding is insufficient or interrupted, and formula is not used as a substitute. In such cases, children may be introduced prematurely to nutrient-poor complementary foods, increasing the risk of malnutrition in both forms. The elevated DBM risk may therefore stem not from the absence of formula alone, but from broader inadequacies in early feeding practices and nutritional care during infancy [34]. The protective effect observed among those introduced to semi-solid foods between six and eight months reinforces WHO recommendations for appropriate complementary feeding timing [35,36].

Maternal education significantly influenced the likelihood of children experiencing the double burden of malnutrition. Those whose mothers had tertiary education were substantially less affected, suggesting that education equips women with the skills to make informed health and nutrition decisions, navigate healthcare systems, and critically assess child feeding recommendations. This supports prior evidence that maternal education enhances not only health literacy but also economic independence and responsiveness to early signs of nutritional distress [37]. Even mothers with only basic education showed marginally reduced odds of having children with DBM, indicating that any formal education can confer benefits. These findings highlight the need to invest in female education as a long-term strategy for improving child nutrition and addressing structural drivers of malnutrition in urban Ghana.

The patterns observed in this study point to deeper systemic and structural imbalances shaping child nutrition in urban Ghana. The double burden of malnutrition does not arise solely from individual choices but is embedded in broader inequalities involving access to nutritious foods, reliable health information, and quality healthcare services. Similar to trends observed across SSA, the coexistence of undernutrition and overweight often affects urban poor populations who face intersecting challenges such as food insecurity, limited maternal education, and constrained living environments [38,39,40]. Interventions must therefore be multifaceted, thus promoting appropriate feeding, encouraging physical activity, and addressing broader social and economic barriers.

This study advances the evidence base by focusing on child-level DBM, a relatively underexplored dimension of malnutrition in Ghana. The integration of behavioral, maternal, and demographic data offers a holistic view of DBM and supports the need for double-duty actions that address under- and overnutrition simultaneously. Public health strategies should prioritize early-life nutrition, enhance maternal support systems, and strengthen local delivery of targeted nutrition and physical activity programs, especially in rapidly urbanizing settings.

## Conclusion

This study presents empirical evidence on the prevalence and determinants of the double burden of malnutrition among under-five children in an urban Ghanaian setting. The identification of key behavioral and socioeconomic factors, including physical inactivity, inadequate breastfeeding practices, early introduction of complementary foods, and limited maternal education, highlights the multifaceted nature of child nutrition challenges in rapidly urbanizing environments. The results reinforce the urgency of adopting integrated, equity-focused public health strategies that address both undernutrition and overnutrition simultaneously. Strengthening maternal education, improving early-life feeding practices, and ensuring access to safe and supportive urban spaces for children are essential components in efforts to reduce DBM in vulnerable populations.

## Data Availability

The datasets are available from the corresponding author upon reasonable request. Data sharing is subject to obtaining ethical clearance from a recognized institutional review board. Interested researchers and institutions should contact the first author, Deborah Balapou, at.

## Limitations

The cross-sectional design limits causal interpretation of associations with DBM. The reliance on convenience sampling at a single tertiary facility may introduce selection bias, as the study population may not be representative of the broader urban under-five population in Ghana. Maternal self-reports of feeding practices and physical activity may also be subject to recall and social desirability bias. Another limitation is the narrow focus on anthropometric indicators; micronutrient deficiencies, food insecurity, and household socioeconomic status were not assessed. These factors could significantly influence nutritional outcomes and should be explored in future research.

## Declarations

### Ethics Approval and Consent

Ethical clearance for this study was obtained from the Komfo Anokye Teaching Hospital Institutional Review Board (KATH/IRB/2023). Written informed consent was obtained from all participating mothers after the purpose and procedures of the study were explained. Participation was voluntary, and confidentiality was ensured through anonymized data collection and secure data storage.

### Consent for Publication

Not applicable.

## Acknowledgments

The authors would like to thank the management and staff of Komfo Anokye Teaching Hospital for their support during data collection. We also appreciate the mothers and children who participated in this study for their time and cooperation. Special thanks go to the research assistants for their dedication and commitment throughout the data collection process.

## Authors’ Contribution

We confirm that this manuscript was solely authored by us. DB, JFAD, and WKAA reviewed literature, contributed to the study design, data collection, data analysis, and manuscript drafting and writing. ROA and EK provided supervision, methodological guidance, and critical revisions of the manuscript. All authors reviewed and approved the final version of the manuscript.

